# Risk factors for rhinitis, allergic conjunctivitis and eczema among schoolchildren in Uganda

**DOI:** 10.1101/2020.06.03.20121251

**Authors:** Harriet Mpairwe, Gyaviira Nkurunungi, Pius Tumwesige, Hellen Akurut, Milly Namutebi, Irene Nambuya, Marble Nnaluwooza, Barbara Apule, Caroline Onen, Tonny Katongole, Emmanuel Niwagaba, Mike Mukasa, Emily L Webb, Alison M Elliott, Neil Pearce

**Affiliations:** Medical Research Council/Uganda Virus Research Institute and London School of Hygiene and Tropical Medicine Uganda Research Unit. Plot 51-59 Nakiwogo Road, Box 49, Entebbe, Uganda; London School of Hygiene and Tropical Medicine. Keppel St, Bloomsbury, London WC1E 7HT, UK

## Abstract

**Background:** The prevalence of allergy-related diseases (ARDs), including rhinitis, allergic conjunctivitis and eczema, is on the increase in Africa and globally. The causes of this increase are not well established.

**Objectives:** To investigate the risk factors for ARDs among schoolchildren in Uganda.

**Methods:** We conducted a secondary data analysis of a large asthma case-control study involving 1,700 schoolchildren, 5-17 years, in urban Uganda. ARDs were defined according to the International Study of Asthma and Allergies in Childhood (ISAAC) questionnaire. Skin prick testing (SPT) was conducted using standard procedures and allergen-specific IgE (asIgE) using ImmunoCAP^®^. We used inverse probability weighting to account for the differences in the sampling fractions in all our analyses.

**Results:** The lifetime prevalence of reported rhinitis, allergic conjunctivitis and eczema was 43.3%, 39.5%, and 13.5%, respectively. There was overlap of ARDs, with 66.3% of 1,193 schoolchildren who reported having ever an ARDs (including asthma) reporting two or more. The important risk factors for ‘rhinitis ever’ were city residence at birth [adjusted odds ratio (95% confidence interval) 1.97 (1.26-3.10) compared to rural]; father’s [2.08 (1.57-2.75)] and mother’s history of allergic disease [2.29 (1.81-2.91)]; frequent de-worming in the last 12 months [1.80 (1.32-2.45), ≥2 versus none]; current high frequency of ‘trucks passing on the street near home’ [1.90 (1.19-3.03), ‘almost all the time’ versus rarely] and positive SPT [1.56 (1.24-1.96)] but not asIgE [1.33 (0.81-2.18)]. The same pattern of risk factors was observed for allergic conjunctivitis and eczema.

**Conclusion:** We found extensive multi-morbidity of, and overlap in the risk factors for, rhinitis, conjunctivitis, and eczema - similar to asthma risk factors - among schoolchildren in urban Uganda. This suggests a similar underlying cause for all ARDs, associated with exposure to urban lifestyles and environment in Uganda. Thus, epidemiological research should investigate causes of all ARDs as one disease entity.

## Introduction

Allergy-related diseases (ARDs) including rhinitis, allergic conjunctivitis and eczema are on the increase globally^1^, ^2^, but the causes of these diseases are generally not established^3^. These chronic recurrent conditions cause significant physical and psychological distress, sleep disturbance and reduced quality of life among people of all ages, but particularly among children^3^, ^4^ The worldwide prevalence is estimated as 42% for rhinitis^2^, 25% for allergic conjunctivitis^4^, and 8% for eczema^5^.

In Africa, and other low and middle income countries (LMICs), the prevalence of these conditions is higher in urban than rural areas^6-9^. Although data on risk factors for ARDs from Africa is scarce, there is evidence to suggest that there may be important differences in risk factors between high income countries (HICs) and LMICs. For example, the International Study of Asthma and Allergies in Childhood (ISAAC) study reported a weaker association between ARDs and allergic sensitisation in LMICs than in HICs^10^. These differences were also supported by results from our own work on risk factors for asthma, another important ARD, among Ugandan schoolchildren^11^. Children born in rural areas had the lowest asthma risk, but this was not associated with exposure to farm animals in early life^11^ as has been found in Europe^12^, ^13^ and North America^14^, ^15^ Childhood asthma was also associated with a higher parental education and social-economic status^11^, in contrast to HICs where asthma is associated with low parental education and social economic status^16^, ^17^ Understanding the risk factors for ARDs in Africa is key to identifying the causes of these conditions and will inform local intervention strategies for prevention and treatment. We therefore undertook a secondary data analysis of a large asthma case-control study involving schoolchildren in urban Uganda, in order to investigate the risk factors for rhinitis, allergic conjunctivitis and eczema.

## Methods

This was a secondary analysis of data from an asthma case-control study^11^. The data that support the findings of this study are available in London School of Hygiene & Tropical Medicine Data Compass at http://datacompass.lshtm.ac.uk/1761/. Data access is restricted due to the presence of potential identifiers^18^. We report our findings according to the STROBE guidelines^19^.

### Study population and enrolment procedures

Schoolchildren, 5-17 years, were enrolled from both primary and secondary schools in an urban area of Wakiso district in central Uganda between May 2015 and July 2017; further details of the study are described elsewhere^11^, ^20^ For each child with asthma (“cases”), two children without a history of asthma symptoms (“controls”) were randomly selected from the class register, using a random number generator programme in STATA (StataCorp, Texas, USA). The parents or guardians of potential participants were contacted to attend a meeting, using invitation cards delivered by the children or telephone calls by the study team. During the meeting, parents/guardians who were interested in having their children participate provided written informed consent. Children eight years or older provided written informed assent.

The study conforms to the standards of the Declaration of Helsinki, and was approved by the Uganda Virus Research Institute Research and Ethics Committee (reference number GC/127/14/09/481), and by the Uganda National Council for Science and Technology (reference number HS 1707).

### Study procedures

Data on rhinitis, allergic conjunctivitis and eczema were collected using the widely used and validated ISAAC questionnaire^21^, which was administered by the study team in either English or Luganda (a language widely understood by the study population). Rhinitis was defined as ‘a problem with sneezing, or a runny, or a blocked nose when you did not have a cold or the flu’; allergic conjunctivitis was defined as ‘recurrent itchy-watery eyes’; eczema was defined as ‘an itchy rash which was coming and going for at least 6 months, and in any of the following places: the folds of the elbows, behind the knees, in front of the ankles, under the buttocks, or around the neck, ears or eyes’. We included an additional question for urticarial rash which was defined as ‘itchy rash associated with wheals (*ebilogologo* in Luganda, a well-known terminology). Questions were answered by either the parents or the participant themselves (for adolescents).

We used the ISAAC environmental questionnaire^22^ to collect data on risk factors for ARDs, and added questions relevant to this setting, such as residence at birth and in the first five years of life [rural or urban (small town or the city Kampala)], and frequency of de-worming in the last 12 months.

We conducted assessments for allergic sensitisation. Skin prick testing (SPT) was conducted using standard procedures^23^ with seven crude allergen extracts *[Dermatophagoides* mix of *D. farinae* and *D. pteronyssnus* (dust mite), *Blomia tropicalis* (dust mite), *Blattella germanica* (cockroach), *Arachis hypogaea* (peanut), cat, pollen mix of weeds, mould mix of *Aspergillus* species; ALK Abello, Hoersholm, Denmark]. Fractional exhaled nitric oxide (FENO) was measured using a handheld device (NoBreath®, Bedfonf Scientific, Maidstone, United Kingdom). We used the manufacturer’s cut-off for children of ≥35 parts per billion. For allergen-specific IgE (asIgE), 200 aliquots of plasma were randomly selected from all participants for testing by ImmunoCAP® (Phadia, Uppsala, Sweden)^24^ using three crude allergen extracts (*D. pteronyssinus, B. germanica* and *Arachis hypogaea*). The standard cut-off for allergic sensitisation of ≥0.35 allergen-specific kilo units per litre (kU_A_/L) was used. Total IgE was also measured using ImmunoCAP®.

Other assessments included the tuberculin skin test (TST) and stool examinations. TST was conducted using standard procedures we have described previously^25^. Examination for helminths was conducted on three stool samples freshly collected on different days, using the Kato Katz method^26^, and these included *Schistosoma mansoni, Trichuris trichuria*, hookworm, and *Ascaris lumbricoides*.

### Statistical considerations

Data were collected on paper questionnaires and double data-entered into OpenClinica open source software version 3.1.4 (OpenClinica LLC and collaborators, Waltham, MA, USA). Cleaned data were transferred to STATA version 15 for analysis. Variables with clinically-relevant standard cut-off points, such as SPT, asIgE, FENO and TST, were analysed as binary variables. Total IgE was analysed as a continuous variable. The variable father’s and mother’s ‘history of allergic disease’ included a history of asthma, rhinitis, conjunctivitis, eczema and any other allergies such as urticaria. We used chi squared tests for comparison of the different risk factors among children with or without a given allergy-related condition.

This was a secondary analysis of data from an asthma case-control study, and therefore it was not a random population sample. The population prevalence of asthma in urban Uganda is approximately 12%^27^, and our asthma case-control study enrolled all identified asthma cases in the population under study [in Wakiso District] and randomly selected twice the number of controls^11^. One might therefore expect that for every 100 children in the source population, there would be 12 asthma cases and 24 non-asthmatic controls (out of 88 non-asthmatics) selected. We therefore assumed that all cases in the source population were sampled, and that the sampling fraction in the non-asthmatics was 24/88 (0.273). We used inverse probability weighting to account for these sampling fractions in all our analyses, i.e. asthma cases received a weight of 1, whereas non-asthmatics received a weight of 3.67 (=1/0.273).

We built each multiple logistic regression model by adding one confounder (identified in literature, and in preliminary analyses of the data) at a time and noted the change in effect side, we stopped adding when there was no change in effect size^28^. Variables that were strongly related (such as mother’s and father’s education, or area of residence at birth and area of residence in the first five years) were not included in the same model in order to minimize problems of collinearity^28^.

In order to investigate whether the results on risk factors for rhinitis, allergic conjunctivitis and eczema were being driven by asthma, we conducted a sub-group analysis among schoolchildren with and without asthma separately, and obtained similar findings. We present results from the weighed analysis.

## Results

For this analysis, we included all 1,700 schoolchildren enrolled in the asthma case-control study^18^. The detailed participant flow diagram has been published previously^11^.

### Prevalence of rhinitis, allergic conjunctivitis and eczema

The lifetime prevalence of reported rhinitis, allergic conjunctivitis and eczema was 43.3%, 39.5%, and 13.5%, respectively, while the prevalence in the last 12 months was 10.1%, 9.1% and 2.3% respectively (Table 1). There was overlap of these ARDs (Table 1); of the 1,193 schoolchildren who reported having ever had ARDs, 791 (66.3%) reported either two, three or four ARDs. The lifetime prevalence of urticarial rash and prevalence in the last 12 months was 31.9% and 1.8%, respectively (Table 1). The overall prevalence of positive skin prick test (to any of seven allergens) at enrolment was 34.7%.

**Table 1:** Weighted prevalence of allergy-related diseases among schoolchildren in Uganda (N=1,700)

| Allergy-related diseases (ARDs) | Ever |  | Last 12 months |  |
| --- | --- | --- | --- | --- |
|  | n | Weighted % | n | Weighted % |
| **1. Overall,** |  |  |  |  |
| Rhinitis | 870 | 43.3 | 216 | 10.1 |
| Allergic conjunctivitis | 772 | 39.5 | 179 | 9.1 |
| Eczema | 265 | 13.5 | 60 | 2.3 |
| Urticarial rash | 572 | 31.9 | 36 | 1.8 |
| **2. ARDs overlap,** |  |  |  |  |
| None (of the four ARDs) | 498 | 38.8 | 959 | 74.8 |
| Asthma only | 60 | 1.3 | 388 | 8.3 |
| Rhinitis only | 171 | 13.3 | 63 | 4.9 |
| Conjunctivitis only | 144 | 11.2 | 59 | 4.6 |
| Eczema only | 27 | 2.1 | 16 | 1.3 |
| 2 ARDs | 396 | 21.6 | 134 | 4.5 |
| 3 ARDs | 308 | 9.9 | 63 | 1.4 |
| 4 ARDs | 87 | 1.8 | 7 | 0.2 |
| **Total** | 1691<sup>a</sup> | 100 | 1689<sup>b</sup> | 100 |

N=number, %=percentage. Missing numbers: <sup>a</sup>=9, <sup>b</sup>=11. Overall, 1,193 (61.2%) reported at least one ARD ever, and 730 (25.2%) in the last 12 months

For the rest of the analysis, the main outcomes are the lifetime history of rhinitis, allergic conjunctivitis, and eczema because of larger numbers compared to the prevalence of these same conditions in the last 12 months.

### Risk factors for rhinitis

Schoolchildren with a lifetime history of rhinitis were more likely than their counterparts without rhinitis to report a father’s [adjusted odds ratio (95% confidence interval), 2.08 (1.57-2.75)]; and mother’s history of allergic disease [2.29 (1.81-2.91)]; residing in the city at birth [1.97 (1.26-3.10)]; having a father with tertiary education [1.37 (1.03-1.80)]; the highest reported frequency of de-worming in the last 12 months [1.80 (1.32-2.45)] and the highest reported frequency of ‘trucks passing on the street near their home’ currently [1.90 (1.19-3.03)] (Table 2). They were more likely to have a positive skin prick test [1.56 (1.24-1.96)] and elevated FENO levels [2.06 (1.57-2.70)], but there were no differences for asIgE and total IgE (Table 2). They were less likely to have a positive tuberculin skin test at enrolment [0.66 (0.44-1.01)] and had lower prevalence of helminths at enrolment [0.70 (0.50-0.97)] (Table 2). Similar results were observed for rhinitis in the last 12 months, albeit wider confidence intervals were observed, due to smaller numbers (Supplementary Table 1).

**Table 2:** Risk factors for rhinitis among schoolchildren in Uganda (N=1,692)

| Characteristics | Rhinitis ever |  | Adj. OR<sup>‡</sup> (95% CI) | P-value |
| --- | --- | --- | --- | --- |
|  | Yes (N=870) | No (N=822) |  |  |
| **Age, years** Mean (SD) | 11.51 (3.12) | 10.65 (3.08) | 1.08 (1.04-1.12) | <0.0001 |
| **Girls** (942) | 511 (58.7) | 431 (52.4) | 1.39 (1.12-1.74) | 0.003 |
| **Father's education** [m=21] |  |  |  |  |
| None/Primary (437) | 198 (22.9) | 239 (29.6) | 1 |  |
| Secondary (595) | 307 (35.6) | 288 (35.6) | 1.14 (0.87-1.50) |  |
| Tertiary (639) | 358 (41.5) | 281 (34.8) | 1.37 (1.03-1.80)<sup>a</sup> | 0.08 |
| **Father's history of allergic disease** [m=149] |  |  |  |  |
| Yes (363) | 242 (30.1) | 121 (16.4) | 2.08 (1.57-2.75) | <0.0001 |
| **Mother's history of allergic disease** [m=126] |  |  |  |  |
| Yes (592) | 384 (46.9) | 208 (27.8) | 2.29 (1.81-2.91) | <0.0001 |
| **Area of residence at the time of the child's birth** [m=1] |  |  |  |  |
| Rural (354) | 159 (18.3) | 195 (23.7) | 1 |  |
| Town (1,188) | 612 (70.3) | 576 (70.2) | 1.22 (0.93-1.60) |  |
| City (149) | 99 (11.4) | 50 (6.1) | 1.97 (1.26-3.10)<sup>b</sup> | 0.01 |
| **Exposure to animals in first five years of life** [m=57] |  |  |  |  |
| Yes (615) | 331 (39.2) | 284 (36.0) | 1.23 (0.98-1.55) | 0.07 |
| **Frequency of 'trucks passing on street near child's home' at enrolment** [m=1] |  |  |  |  |
| Rarely (944) | 442 (50.9) | 501 (61.0) | 1 |  |
| Frequently (624) | 346 (39.8) | 278 (33.9) | 1.42 (1.13-1.79) |  |
| Almost all the time (123) | 81 (9.3) | 42 (5.1) | 1.90 (1.19-3.03)<sup>c</sup> | 0.001 |
| **Use of de-worming medication in last 12 months** [m=1] |  |  |  |  |
| None (578) | 276 (31.8) | 302 (36.8) | 1 |  |
| Once (732) | 345 (39.7) | 387 (47.1) | 0.85 (0.67-1.09) |  |
| ≥Twice (380) | 248 (28.5) | 132 (16.1) | 1.80 (1.32-2.45)<sup>d</sup> | <0.0001 |
| **Any helminth infection at enrolment** [m=130] |  |  |  |  |
| Yes (212) | 90 (11.2) | 122 (16.0) | 0.70 (0.50-0.97) | 0.03 |
| **Tuberculin skin test induration at enrolment** [N=960] |  |  |  |  |
| Positive (≥10mm) (143) | 66 (13.4) | 77 (16.4) | 0.66 (0.44-1.01) | 0.05 |
| **Skin Prick Test response to any of 7 allergens at enrolment** [m=36] |  |  |  |  |
| Positive (≥3mm) (657) | 392 (45.8) | 265 (33.1) | 1.56 (1.24-1.96) | <0.0001 |
| **Fractional exhaled nitric oxide levels at enrolment** [m=80] |  |  |  |  |
| Elevated (≥35ppb) (381) | 254 (30.2) | 127 (16.4) | 2.06 (1.57-2.70) | <0.0001 |
| **Allergen-specific IgE to any of 3 allergens at enrolment** (N=397)<sup>§</sup> |  |  |  |  |
| Atopic (≥0.35 kU<sub>A</sub>/L) (251) | 142 (66.4) | 109 (59.6) | 1.33 (0.81-2.18) | 0.26 |
| **Total IgE** (N=398) Median (95%CI) kU/L | 414.8 (281.1-537.5) | 304.3 (213.6-359.3) | 1.00002 (0.99983-1.00020) | 0.87 |

N=number; Adj. OR=adjusted odds ratio; CI=confidence interval; SD=standard deviation; m=missing; Columns 2 and 3 represent number (%), unless stated otherwise. <sup>‡</sup>Adjusted for child's age, sex, area of residence at birth and father's education. Test for trend p-value: <sup>a</sup>=0.03; <sup>b</sup>=0.006; <sup>c</sup><0.0001; <sup>d</sup>=0.002. <sup>§</sup> ImmunoCAP®, cut-off ≥0.35 kU<sub>A</sub>/L.

### Risk factors for allergic conjunctivitis

Schoolchildren with a lifetime history of allergic conjunctivitis were more likely than their counterparts without allergic conjunctivitis to report a father’s [2.06 (1.56-2.71)] and mother’s history of allergic disease [1.57 (1.24-1.98)]; residing in the city at birth [1.69 (1.09-2.64)]; being exposed to farm animals in early life [1.33 (1.05-1.67)]; having a father with tertiary education [1.35 (1.03-1.79)]; the highest reported frequency of de-worming in last 12 months [1.94 (1.44-2.63)] and the highest reported frequency of ‘trucks passing on the street near their home’ currently [1.60 (1.03-2.48)] (Table 3).

**Table 3:** Risk factors for allergic conjunctivitis among schoolchildren in Uganda (N=1,691)

| Characteristics | Conjunctivitis ever |  | Adj. OR<sup>‡</sup> (95% CI) | P-value |
| --- | --- | --- | --- | --- |
|  | Yes (N=772) | No (N=919) |  |  |
| **Age, years Mean (SD)** | 11.45 (3.10) | 10.80 (3.12) | 1.06 (1.02-1.10) | 0.002 |
| **Girls (942)** | 430 (55.7) | 512 (55.7) | 1.02 (0.82-1.27) | 0.83 |
| **Father's education [m=20]** |  |  |  |  |
| None/Primary (437) | 172 (22.4) | 265 (29.3) | 1 |  |
| Secondary (595) | 284 (37.1) | 311 (34.4) | 1.28 (0.97-1.69) |  |
| Tertiary (639) | 310 (40.5) | 329 (36.3) | 1.35 (1.03-1.79)<sup>a</sup> | 0.08 |
| **Father's history of allergic disease [m=148]** |  |  |  |  |
| Yes (363) | 222 (31.2) | 141 (17.0) | 2.06 (1.56-2.71) | <0.0001 |
| **Mother's history of allergic disease [m=125]** |  |  |  |  |
| Yes (592) | 315 (43.4) | 277 (32.9) | 1.57 (1.24-1.98) | <0.0001 |
| **Area of residence at the time of the child's birth** |  |  |  |  |
| Rural (354) | 158 (20.5) | 196 (21.3) | 1 |  |
| Town (1,188) | 523 (67.7) | 665 (72.4) | 0.89 (0.68-1.16) |  |
| City (149) | 91 (11.8) | 58 (6.3) | 1.69 (1.09-2.64)<sup>b</sup> | 0.006 |
| **Exposure to animals in first five years of life [m=58]** |  |  |  |  |
| Yes (615) | 296 (39.7) | 319 (35.9) | 1.33 (1.05-1.67) | 0.02 |
| **Frequency of 'trucks passing on street near child's home' at enrolment** |  |  |  |  |
| Rarely (944) | 411 (53.2) | 533 (58.0) | 1 |  |
| Frequently (624) | 290 (37.6) | 334 (36.3) | 1.06 (0.84-1.34) |  |
| Almost all the time (123) | 71 (9.2) | 53 (5.7) | 1.60 (1.03-2.48)<sup>c</sup> | 0.05 |
| **Use of de-worming medication in last 12months [m=1]** |  |  |  |  |
| None (578) | 228 (29.6) | 350 (38.1) | 1 |  |
| Once (732) | 320 (41.5) | 412 (44.8) | 1.09 (0.85-1.40) |  |
| ≥Twice (380) | 223 (28.9) | 157 (17.1) | 1.94 (1.44-2.63)<sup>d</sup> | <0.0001 |
| **Any helminth infection at enrolment [m=129]** |  |  |  |  |
| Yes (212) | 87 (12.1) | 125 (14.8) | 0.88 (0.63-1.22) | 0.44 |
| **Tuberculin skin test induration at enrolment [N=960]** |  |  |  |  |
| Positive (≥10mm) (143) | 58 (13.9) | 85 (15.6) | 0.68 (0.45-1.05) | 0.08 |
| **Skin Prick Test response to any of 7 allergens at enrolment [m=35]** |  |  |  |  |
| Positive (≥3mm) (657) | 368 (48.9) | 289 (32.0) | 1.76 (1.40-2.21) | <0.0001 |
| **Fractional exhaled nitric oxide levels at enrolment [m=79]** |  |  |  |  |
| Elevated (≥35ppb) (381) | 227 (30.2) | 154 (17.9) | 1.57 (1.20-2.05) | 0.001 |
| **Allergen-specific IgE at enrolment (N=397)<sup>§</sup>** |  |  |  |  |
| Atopic (≥0.35 kU<sub>A</sub>/L) (251) | 136 (67.3) | 155 (59.0) | 1.22 (0.75-1.98) | 0.41 |
| **Total IgE at enrolment (N=398)** |  |  |  |  |
| Median (95%CI) kU/L | 335.2 (250.7-457.6) | 335.8 (270.9-420.0) | 0.9999 (0.9997-1.0001) | 0.28 |

N=number; Adj. OR=adjusted odds ratio; CI=confidence interval; SD=standard deviation; m=missing; Columns 2 and 3 represent number (%), unless stated otherwise. <sup>‡</sup>Adjusted for child's age, sex, area of residence at birth and father's education. Test for trend p-value: <sup>a</sup>=0.03; <sup>b</sup>=0.22; <sup>c</sup>=0.09; <sup>d</sup><0.0001 <sup>§</sup>ImmunoCAP®, cut-off ≥0.35 kU<sub>A</sub>/L.

They were more likely to have a positive SPT [1.76 (1.40-2.21)] and elevated FENO levels [1.57 (1.20-2.05)], but there was no difference for asIgE and total IgE (Table 3). Similar results were observed for allergic conjunctivitis in the last 12 months, albeit with wider confidence intervals due to much smaller numbers.

### Risk factors for eczema

Children with a lifetime history of eczema were more likely than their counterparts without eczema to report a father’s [2.19 (1.56-3.09)] and mother’s history of allergic disease [1.94 (1.42-2.67)]; residing in the city at birth [1.55 (0.91-2.63)]; the highest reported frequency of de-worming in last 12 months [1.63 (1.10-2.43)] and the highest reported frequency of ‘trucks passing on the street near their home’ currently [1.94 (1.14-3.32)], but there were no differences for SPT, asIgE and total IgE (Table 4). Numbers for eczema in the last 12 months were small.

**Table 4:**
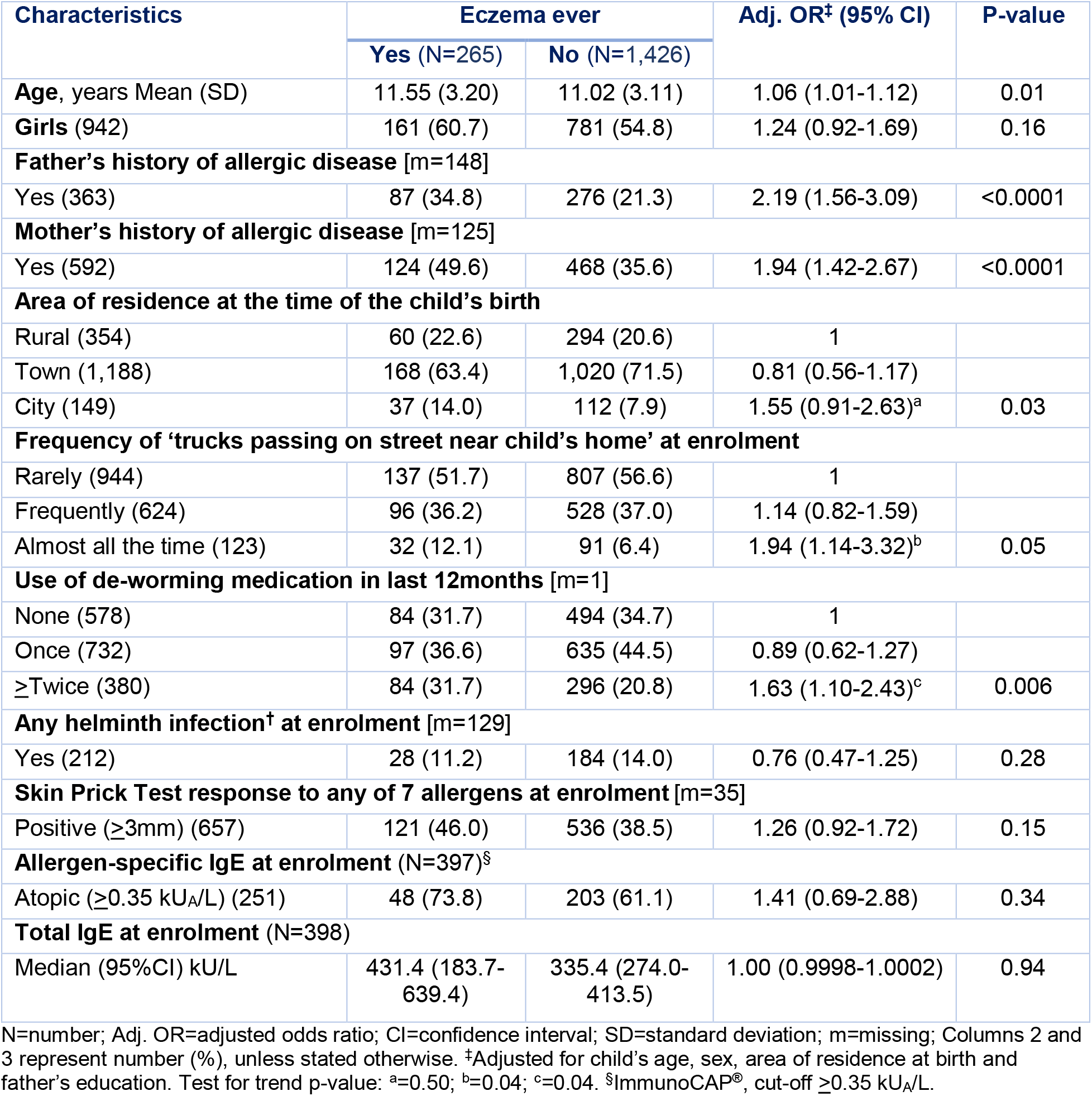
Risk factors for eczema among schoolchildren in Uganda (N=1,691)

| Characteristics | Eczema ever |  | Adj. OR<sup>‡</sup> (95% CI) | P-value |
| --- | --- | --- | --- | --- |
|  | Yes (N=265) | No (N=1,426) |  |  |
| **Age**, years Mean (SD) | 11.55 (3.20) | 11.02 (3.11) | 1.06 (1.01-1.12) | 0.01 |
| **Girls** (942) | 161 (60.7) | 781 (54.8) | 1.24 (0.92-1.69) | 0.16 |
| **Father's history of allergic disease** [m=148] |  |  |  |  |
| Yes (363) | 87 (34.8) | 276 (21.3) | 2.19 (1.56-3.09) | <0.0001 |
| **Mother's history of allergic disease** [m=125] |  |  |  |  |
| Yes (592) | 124 (49.6) | 468 (35.6) | 1.94 (1.42-2.67) | <0.0001 |
| **Area of residence at the time of the child's birth** |  |  |  |  |
| Rural (354) | 60 (22.6) | 294 (20.6) | 1 |  |
| Town (1,188) | 168 (63.4) | 1,020 (71.5) | 0.81 (0.56-1.17) |  |
| City (149) | 37 (14.0) | 112 (7.9) | 1.55 (0.91-2.63)<sup>a</sup> | 0.03 |
| **Frequency of 'trucks passing on street near child's home' at enrolment** |  |  |  |  |
| Rarely (944) | 137 (51.7) | 807 (56.6) | 1 |  |
| Frequently (624) | 96 (36.2) | 528 (37.0) | 1.14 (0.82-1.59) |  |
| Almost all the time (123) | 32 (12.1) | 91 (6.4) | 1.94 (1.14-3.32)<sup>b</sup> | 0.05 |
| **Use of de-worming medication in last 12months** [m=1] |  |  |  |  |
| None (578) | 84 (31.7) | 494 (34.7) | 1 |  |
| Once (732) | 97 (36.6) | 635 (44.5) | 0.89 (0.62-1.27) |  |
| ≥Twice (380) | 84 (31.7) | 296 (20.8) | 1.63 (1.10-2.43)<sup>c</sup> | 0.006 |
| **Any helminth infection<sup>†</sup> at enrolment** [m=129] |  |  |  |  |
| Yes (212) | 28 (11.2) | 184 (14.0) | 0.76 (0.47-1.25) | 0.28 |
| **Skin Prick Test response to any of 7 allergens at enrolment** [m=35] |  |  |  |  |
| Positive (≥3mm) (657) | 121 (46.0) | 536 (38.5) | 1.26 (0.92-1.72) | 0.15 |
| **Allergen-specific IgE at enrolment** (N=397)<sup>§</sup> |  |  |  |  |
| Atopic (≥0.35 kU<sub>A</sub>/L) (251) | 48 (73.8) | 203 (61.1) | 1.41 (0.69-2.88) | 0.34 |
| **Total IgE at enrolment** (N=398) |  |  |  |  |
| Median (95%CI) kU/L | 431.4 (183.7-639.4) | 335.4 (274.0-413.5) | 1.00 (0.9998-1.0002) | 0.94 |

N=number; Adj. OR=adjusted odds ratio; CI=confidence interval; SD=standard deviation; m=missing; Columns 2 and 3 represent number (%), unless stated otherwise. <sup>‡</sup>Adjusted for child's age, sex, area of residence at birth and father's education. Test for trend p-value: <sup>a</sup>=0.50; <sup>b</sup>=0.04; <sup>c</sup>=0.04. <sup>§</sup>ImmunoCAP®, cut-off ≥0.35 kU<sub>A</sub>/L.

### Risk factors for ARDs combined

Because of the high rate of multi-morbidity of ARDs (Table 1) and the high overlap of risk factors for these conditions (Tables 2-4), we created a new variable that combined all the ARDs (including asthma), and compared children with any ARD to children who did not report any ARDs. We found the risk factors for ‘any ARD ever’ to include a father’s [2.64 (1.89-3.70)] and mother’s history of allergic disease [2.21 (1.70-2.86)]; city residence at birth [2.12 (1.26-3.57)]; exposure to farm animals in first five years of life [1.41 (1.11-1.80)]; the highest reported frequency of de-worming in the last 12 months [2.12 (1.51-2.99)] and ‘trucks passing on the street near home’ currently [2.18 (1.25-3.78)]; positive SPT [1.91 (1.49-2.46)] and elevated FENO [1.92 (1.40-2.64)], but not asIgE or total IgE (Table 5). These results were similar for ARDs in the last 12 months, except this time and association asIgE was statistically significant [1.85 (1.14-3.01)] (Supplementary Table 2). Similar results were observed for risk factors for ARDs among non-asthma controls only (Supplementary Table 3).

**Table 5:** Risk factors for having ever had any of rhinitis, conjunctivitis, eczema and asthma among Ugandan schoolchildren (N=1,693)

| Characteristics | Allergy-related diseases – ever |  | Adj. OR<sup>‡</sup> (95% CI) | P-value |
| --- | --- | --- | --- | --- |
|  | Yes (N=1,195) | No (N=498) |  |  |
| **Age**, years Mean (SD) | 11.29 | 10.64 | 1.06 (1.02-1.10) | 0.003 |
| **Girls** (942) | 680 (56.9) | 262 (52.6) | 1.28 (1.02-1.61) | 0.03 |
| **Father's history of allergic disease** [m=150] |  |  |  |  |
| Yes (363) | 313 (28.5) | 50 (11.2) | 2.64 (1.89-3.70) | <0.0001 |
| **Mother's history of allergic disease** [m=127] |  |  |  |  |
| Yes (592) | 483 (43.1) | 109 (24.4) | 2.21 (1.70-2.86) | <0.0001 |
| **Area of residence at the time of the child's birth** [m=1] |  |  |  |  |
| Rural (355) | 234 (19.6) | 121 (24.3) | 1 |  |
| Town (1,188) | 835 (69.9) | 353 (70.9) | 1.06 (0.81-1.40) |  |
| City (149) | 125 (10.5) | 24 (4.8) | 2.12 (1.26-3.57)<sup>a</sup> | 0.01 |
| **Exposure to animals in first five years of life** [m=60] |  |  |  |  |
| Yes (615) | 454 (39.2) | 161 (33.8) | 1.41 (1.11-1.80) | 0.005 |
| **Frequency of 'trucks passing on street near child's home' at enrolment** [m=1] |  |  |  |  |
| Rarely (945) | 626 (52.4) | 319 (64.1) | 1 |  |
| Frequently (624) | 464 (38.9) | 160 (32.1) | 1.40 (1.10-1.78) |  |
| Almost all the time (123) | 104 (8.7) | 19 (3.8) | 2.18 (1.25-3.78)<sup>b</sup> | 0.001 |
| **Use of de-worming medication in last 12 months** [m=3] |  |  |  |  |
| None (578) | 384 (32.2) | 194 (39.0) | 1 |  |
| Once (732) | 491 (41.2) | 241 (48.4) | 0.95 (0.75-1.22) |  |
| ≥Twice (380) | 317 (26.6) | 63 (12.6) | 2.12 (1.51-2.99)<sup>c</sup> | <0.0001 |
| **Any helminth infection at enrolment** [m=130] |  |  |  |  |
| Yes (212) | 133 (12.0) | 79 (17.3) | 0.73 (0.53-1.02) | 0.06 |
| **Tuberculin skin test induration at enrolment** [N=961] |  |  |  |  |
| Positive (≥10mm) (143) | 88 (13.1) | 55 (19.0) | 0.60 (0.39-0.90) | 0.01 |
| **Skin Prick Test response to any of 7 allergens at enrolment** [m=36] |  |  |  |  |
| Positive (≥3mm) (657) | 529 (45.2) | 128 (26.3) | 1.91 (1.49-2.46) | <0.0001 |
| **Fractional exhaled nitric oxide levels at enrolment** [m=80] |  |  |  |  |
| Elevated (≥35ppb) (381) | 321 (27.7) | 60 (13.2) | 1.92 (1.40-2.64) | <0.0001 |
| **Allergen-specific IgE at enrolment** (N=398)<sup>§</sup> |  |  |  |  |
| Atopic (≥0.35 kU<sub>A</sub>/L) (252) | 203 (67.2) | 49 (51.0) | 1.54 (0.92-2.56) | 0.10 |
| **Total IgE** (N=398) Median (95%CI) kU/L | 348.2 (279.9-465.3) | 310.0 (187.6-381.5) | 0.9998 (0.9996-1.0001) | 0.22 |

N=number; Adj. OR=adjusted odds ratio; CI=confidence interval; SD=standard deviation; m=missing; Columns 2 and 3 represent number (%), unless stated otherwise. <sup>‡</sup>Adjusted for child's age, sex, area of residence at birth and father's education. Test for trend p-value: <sup>a</sup>=0.02; <sup>b</sup><0.0001; <sup>c</sup><0.0001. <sup>§</sup> ImmunoCAP, cut-off ≥0.35 kU<sub>A</sub>/L.

### Risk factors for positive skin prick test

Children with positive SPT were more likely than SPT-negative children to report having a father [1.36 (1.02-1.81)] and/or a mother with tertiary education [1.72 (1.29-2.30)]; a father’s [1.41 (1.06-1.87)] and mother’s history of allergic disease [1.28 (1.00-1.63)]; residing in the city at birth [1.83 (1.17-2.86)]; the highest reported frequency of ‘trucks passing on the street near their home’ currently [2.26 (1.45-3.55)]; and were more likely to have elevated FENO [4.38 (3.29-5.82)], allergen-specific [25.91 (10.53-63.80)] and total IgE (Table 6). Similar results were observed in the sub-group analysis of only children without asthma.

**Table 6:** Risk factors for skin prick test reactivity among schoolchildren in Uganda (N=1,661)

| Characteristics | Skin prick test<sup>#</sup> |  | Adj. OR<sup>‡</sup> (95% CI) | P-value |
| --- | --- | --- | --- | --- |
|  | Pos. (N=658) | Neg. (N=1,003) |  |  |
| Age, years Mean (SD) | 11.18 (3.10) | 11.08 (3.14) | 1.03 (1.00-1.07) | 0.08 |
| Girls (930) | 308 (46.8) | 622 (62.0) | 0.57 (0.45-0.71) | <0.0001 |
| **Father's highest education level [m=25]** |  |  |  |  |
| None/Primary (425) | 147 (22.5) | 278 (28.3) | 1 |  |
| Secondary (585) | 215 (32.9) | 370 (37.6) | 0.92 (0.68-1.23) |  |
| Tertiary (626) | 291 (44.6) | 335 (34.1) | 1.36 (1.02-1.81)<sup>a</sup> | 0.01 |
| **Mother's highest education level [m=26]** |  |  |  |  |
| None/Primary (569) | 186 (28.5) | 383 (39.0) | 1 |  |
| Secondary (590) | 229 (35.1) | 361 (36.8) | 1.26 (0.96-1.64) |  |
| Tertiary (476) | 238 (36.4) | 238 (24.2) | 1.72 (1.29-2.30)<sup>b</sup> | 0.001 |
| **Father's history of allergic disease [m=125]** |  |  |  |  |
| Yes (356) | 178 (29.2) | 178 (19.7) | 1.41 (1.06-1.87) | 0.02 |
| **Mother's history of allergic disease [m=95]** |  |  |  |  |
| Yes (577) | 264 (39.7) | 331 (36.1) | 1.28 (1.00-1.63) | 0.05 |
| **Area of residence at the time of the child's birth [m=4]** |  |  |  |  |
| Rural (348) | 113 (17.2) | 235 (23.5) | 1 |  |
| Town (1,164) | 467 (71.1) | 697 (69.7) | 1.19 (0.89-1.59) |  |
| City (145) | 77 (11.7) | 68 (6.8) | 1.83 (1.17-2.86)<sup>c</sup> | 0.03 |
| **Frequency of 'trucks passing on street near child's home' at enrolment [m=4]** |  |  |  |  |
| Rarely (923) | 340 (51.8) | 583 (58.3) | 1 |  |
| Frequently (615) | 255 (38.8) | 360 (36.0) | 1.21 (0.95-1.54) |  |
| Almost all the time (119) | 62 (9.4) | 57 (5.7) | 2.26 (1.45-3.55)<sup>d</sup> | 0.001 |
| **Use of de-worming medication in last 12months [m=6]** |  |  |  |  |
| None (572) | 221 (33.6) | 351 (35.2) | 1 |  |
| Once (713) | 281 (42.8) | 432 (43.3) | 0.97 (0.75-1.25) |  |
| ≥Twice (370) | 155 (23.6) | 215 (21.5) | 0.87 (0.64-1.19)<sup>e</sup> | 0.68 |
| **Any helminth infection<sup>†</sup> at enrolment [m=119]** |  |  |  |  |
| Yes (211) | 73 (11.9) | 138 (14.8) | 0.76 (0.54-1.08) | 0.13 |
| **Fractional exhaled nitric oxide levels at enrolment [m=74]** |  |  |  |  |
| Elevated (≥35ppb) (380) | 261 (41.3) | 119 (12.5) | 4.38 (3.29-5.82) | <0.0001 |
| **Allergen-specific IgE at enrolment (N=395)<sup>§</sup>** |  |  |  |  |
| Atopic (≥0.35 kU<sub>A</sub>/L) (251) | 156 (94.5) | 95 (41.3) | 25.91 (10.53-63.80) | <0.0001 |
| **Total IgE at enrolment (N=396)** |  |  |  |  |
| Median (95%CI) kU/L | 733.2 (587.6-892.7) | 179.0 (123.1-238.7) | 1.0004 (1.0001-1.0008) | 0.01 |

N=number; Pos=positive; Neg=Negative; Adj. OR=adjusted odds ratio; CI=confidence interval; SD=standard deviation; m=missing; Columns 2 and 3 represent number (%), unless stated otherwise. <sup>#</sup>whole allergen extracts of *Blomia tropicalis*, Dermatophagoides mix, cockroach, peanut, cat, weeds pollen mix, and mould mix. <sup>†</sup>Adjusted for child's age, sex, area of residence at birth and father's education. Test for trend p-value: <sup>a</sup>=0.03; <sup>b</sup>=<0.0001; <sup>c</sup>=0.02; <sup>d</sup>=0.001; <sup>e</sup>=0.43. <sup>§</sup>3 allergen extracts (Dermatophagoides, cockroach and peanut), using ImmunoCAP<sup>®</sup> on a random sample of 400; standard cut-off ≥0.35 kU<sub>A</sub>/L.

For urticarial rash, the statistically significant risk factors were a father’s and mother’s history of allergic disease, reported increased frequency of de-worming and of ‘trucks passing on the street near home’ in the last 12 months, but not SPT, asIgE and total IgE (Supplementary Table 4).

Other potential factors, including maternal smoking during pregnancy (3%), current smoking by anyone in the household (including the child, 11%), and breastfeeding history (74% of children breastfed to more than one year) were not associated with ARDs.

## Discussion

We found extensive multi-morbidity and substantial overlap of the risk factors for rhinitis, allergic conjunctivitis, eczema, urticarial rash and atopic sensitisation among schoolchildren in urban Uganda. The most consistent risk factors included parental history of allergic disease, city residence at birth, current proximity to a busy road, frequent de-worming, positive SPT and elevated FENO, but not allergen specific-IgE or total IgE to crude allergen extracts.

We found that children born in the city (a proxy for mother’s residence during pregnancy) were at a higher risk of ARDs than their counterparts born in the village. Of note, the schools were situated in an urban setting, and most of the schoolchildren enrolled were in the ‘day section’ of school and commuted daily within the study area (a predominantly urban setting). Thus current residence was considered as reasonably uniform, and urban. This highlights the importance of exposures in early life and is consistent with the observation that the prevalence of ARDs is higher among children in urban than rural areas in Africa^29^ and other LMICs^9^ This observation is also consistent with studies from Europe and North America that report a lower risk of ARDs among children raised on farms (which are predominantly in rural areas), and this has been attributed to early life exposure to farm animals^12^, ^30^ However, the association between reported early life exposure to farm animals and ARDs was positive. This implies that the observed ‘protective rural effect’ in this setting could not be explained by exposure to farm animals, consistent with studies from other developing countries^31^, ^32^

We found a positive association between ARDs and positive SPT responses and with elevated FENO, which is consistent with underlying allergic inflammation that is common to these conditions^3^, ^4^ However, there was a lack of association between asIgE (to three crude allergen extracts) and rhinitis, conjunctivitis and eczema, despite the standard asIgE assessments and cut-off points used, and a strong association between asIgE and SPT reactivity. This lack of association may be explained, at least in part, by high levels of cross-reactive IgE to other environmental allergens, particularly to cross-reactive carbohydrate determinants, as reported by studies in Ghana^33^. Indeed, our recent work in Uganda showed that associations between asIgE or SPT sensitization and clinical allergy outcomes were weak among participants from rural, compared to urban, settings^34^. Rural residents a higher prevalence of helminths - an important source of ‘environmental’ antigens^34^. Our observations are consistent with the ISAAC study, which reported a weaker association between ARDs and atopic sensitisation in LMICs than in HICs^10^. This weak/lack of association between ARDs and asIgE has important implications for diagnosis and treatment of these conditions in this setting: what proportion of these conditions are *allergic*, and would immunotherapy or biologicals treat as effectively as in HICs? To answer this question would require the use of component-resolved diagnosis (using purified single protein allergens) to determine IgE sensitisation, with high cost implications that may be prohibitive for routine use in this setting.

The participant characteristics that were consistent across all ARDs were the reported high frequency of ‘trucks on the street near home’ and reported frequent use of de-worming medication. We think ‘trucks near home’ is a proxy for proximity to a busy road, and this has been previously found to be positively associated with ARDs^35^, ^36^ and asthma morbidity^37^, and this is probably related to increased pollution. With regards to the reported increased frequency of deworming among children with the different ARDs including asthma^11^, we have no ready explanation. In Uganda, schoolchildren routinely receive mass drug administration with albendazole once a year, but in this study, children with ARDs were more likely to report being dewormed twice or more in the last one year. There was a trend towards low prevalence of helminths among children with ARDs but this was statistically significant for only ‘rhinitis ever’. The inverse association between rhinitis and the tuberculin skin test has been reported previously in South Africa^38^.

A limitation of the study was that we performed standard tests for asIgE and total IgE on a randomly selected 400 schoolchildren, not the entire sample size of 1,700, limiting our power to detect weaker associations with rhinitis, conjunctivitis and eczema (we had good power to detect strong associations). The average age of the participants was 10 years and as such, we could not obtain detailed information on some exposures in early life. Nevertheless, we highlight the importance of environmental exposures in early life in increasing the risk of ARDs in the urban setting.

The strength of this study is the large sample size and the weighed analysis, which make our findings generalisable to schoolchildren in urban areas in Uganda, and probably in sub-Saharan Africa. The risk factors for rhinitis, conjunctivitis, eczema and atopic sensitisation (SPT) were similar to the risk factors for asthma reported in our earlier work^11^, even in the subgroup analysis of children without asthma. Given the similarity in risk factors, and the extensive overlap of these ARDs among children, it is possible that these conditions have the same underlying cause and as such, should be investigated as one entity in population studies. This hypothesis has also been suggested by other authors^39^, and is supported by studies that show the role of shared genetic and environmental factors^40^. The specific shared early life lifestyle and environmental risk factors have not been identified, but there are important differences with findings from HICs: in this setting, there is possibly a reduced role of asIgE and exposure to farm animals, an increased risk associated with higher parental education and socio-economic status^41^, and urbanisation^7^. Investigating these differences will increase our understanding of underlying causes of all allergy-related diseases.

## Conclusion

We found extensive multi-morbidity of, and overlap in the risk factors for, rhinitis, conjunctivitis, and eczema - similar to asthma risk factors - among schoolchildren in urban Uganda. This suggests a similar underlying cause for all ARDs, associated with early-life exposure to urban lifestyles and environment in Uganda. The rapid urbanisation and population growth in Africa provides an excellent opportunity to identify the specific cause. This calls for epidemiological research to investigate the causes of ARDs in urban Africa, as one disease entity.

## Data Availability

The data that support the findings of this study are available in London School of Hygiene & Tropical Medicine Data Compass at http://datacompass.lshtm.ac.uk/1761/. Data access is restricted due to the presence of potential identifiers.

## Acknowledgements

Many thanks to the study participants, their parents, guardians and teachers for their enthusiasm and support during this study. We acknowledge the contribution of Ronald van Ree from Amsterdam University Medical Centers (AMC), The Netherlands, who worked with GN on the asIgE ImmunoCAP® assays. This study was funded by Wellcome (reference no. 102512 Training fellowship to HM; 095778 Senior fellowship to AE), European Research Council (reference no. 668954 to NP), African Partnership for Chronic Disease Research (to GN) and UK Medical Research Council (reference no. MR/K012126/1 to EW).

---

**Supplementary Table 1:**
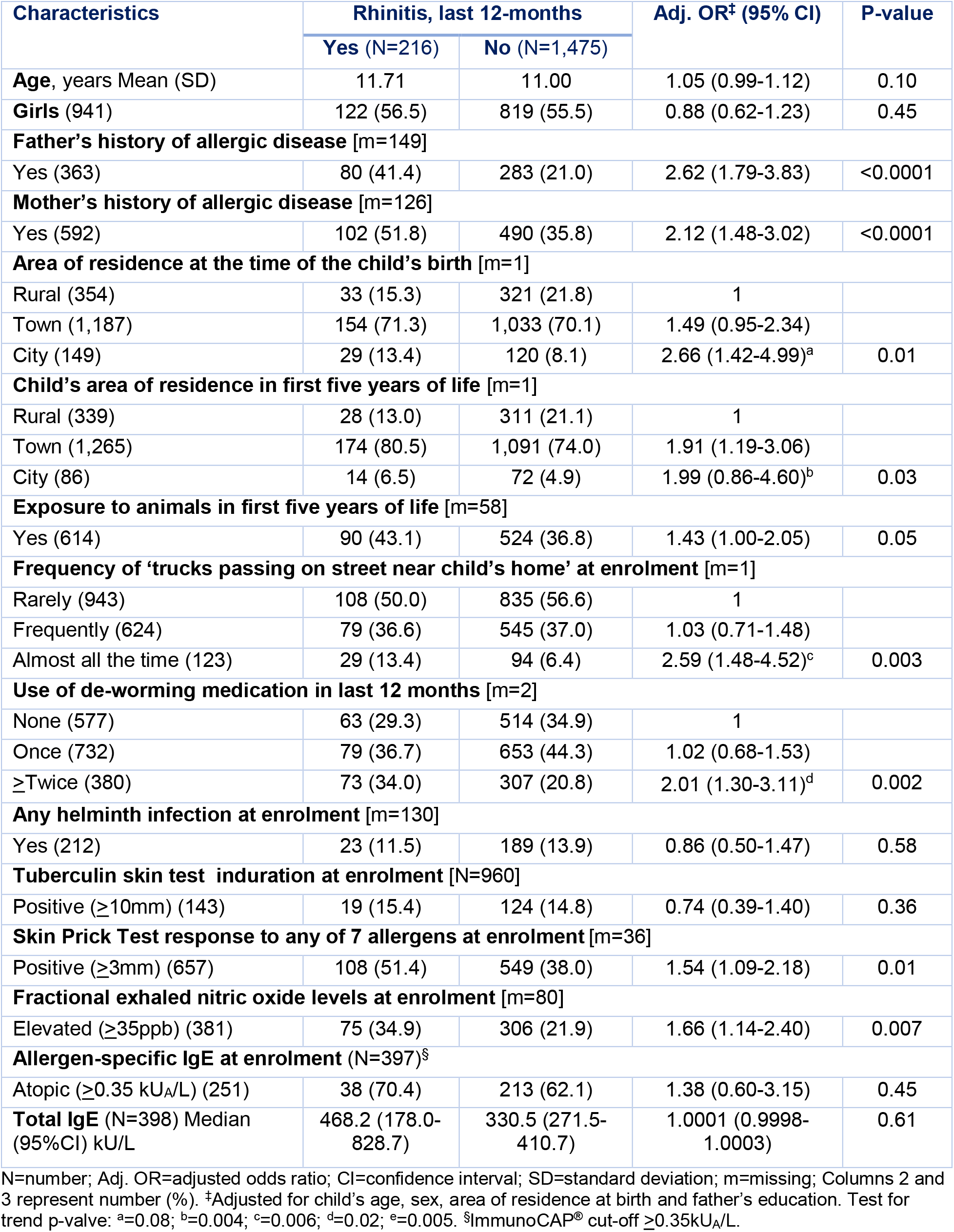
Risk factors for current rhinitis among Ugandan schoolchildren (N=1,691)

| Characteristics | Rhinitis, last 12-months |  | Adj. OR* (95% CI) | P-value |
| --- | --- | --- | --- | --- |
|  | Yes (N=216) | No (N=1,475) |  |  |
| **Age, years Mean (SD)** | 11.71 | 11.00 | 1.05 (0.99-1.12) | 0.10 |
| **Girls (941)** | 122 (56.5) | 819 (55.5) | 0.88 (0.62-1.23) | 0.45 |
| **Father's history of allergic disease [m=149]** |  |  |  |  |
| Yes (363) | 80 (41.4) | 283 (21.0) | 2.62 (1.79-3.83) | <0.0001 |
| **Mother's history of allergic disease [m=126]** |  |  |  |  |
| Yes (592) | 102 (51.8) | 490 (35.8) | 2.12 (1.48-3.02) | <0.0001 |
| **Area of residence at the time of the child's birth [m=1]** |  |  |  |  |
| Rural (354) | 33 (15.3) | 321 (21.8) | 1 |  |
| Town (1,187) | 154 (71.3) | 1,033 (70.1) | 1.49 (0.95-2.34) |  |
| City (149) | 29 (13.4) | 120 (8.1) | 2.66 (1.42-4.99)<sup>a</sup> | 0.01 |
| **Child's area of residence in first five years of life [m=1]** |  |  |  |  |
| Rural (339) | 28 (13.0) | 311 (21.1) | 1 |  |
| Town (1,265) | 174 (80.5) | 1,091 (74.0) | 1.91 (1.19-3.06) |  |
| City (86) | 14 (6.5) | 72 (4.9) | 1.99 (0.86-4.60)<sup>b</sup> | 0.03 |
| **Exposure to animals in first five years of life [m=58]** |  |  |  |  |
| Yes (614) | 90 (43.1) | 524 (36.8) | 1.43 (1.00-2.05) | 0.05 |
| **Frequency of 'trucks passing on street near child's home' at enrolment [m=1]** |  |  |  |  |
| Rarely (943) | 108 (50.0) | 835 (56.6) | 1 |  |
| Frequently (624) | 79 (36.6) | 545 (37.0) | 1.03 (0.71-1.48) |  |
| Almost all the time (123) | 29 (13.4) | 94 (6.4) | 2.59 (1.48-4.52)<sup>c</sup> | 0.003 |
| **Use of de-worming medication in last 12 months [m=2]** |  |  |  |  |
| None (577) | 63 (29.3) | 514 (34.9) | 1 |  |
| Once (732) | 79 (36.7) | 653 (44.3) | 1.02 (0.68-1.53) |  |
| ≥Twice (380) | 73 (34.0) | 307 (20.8) | 2.01 (1.30-3.11)<sup>d</sup> | 0.002 |
| **Any helminth infection at enrolment [m=130]** |  |  |  |  |
| Yes (212) | 23 (11.5) | 189 (13.9) | 0.86 (0.50-1.47) | 0.58 |
| **Tuberculin skin test induration at enrolment [N=960]** |  |  |  |  |
| Positive (≥10mm) (143) | 19 (15.4) | 124 (14.8) | 0.74 (0.39-1.40) | 0.36 |
| **Skin Prick Test response to any of 7 allergens at enrolment [m=36]** |  |  |  |  |
| Positive (≥3mm) (657) | 108 (51.4) | 549 (38.0) | 1.54 (1.09-2.18) | 0.01 |
| **Fractional exhaled nitric oxide levels at enrolment [m=80]** |  |  |  |  |
| Elevated (≥35ppb) (381) | 75 (34.9) | 306 (21.9) | 1.66 (1.14-2.40) | 0.007 |
| **Allergen-specific IgE at enrolment (N=397)<sup>§</sup>** |  |  |  |  |
| Atopic (≥0.35 kU<sub>A</sub>/L) (251) | 38 (70.4) | 213 (62.1) | 1.38 (0.60-3.15) | 0.45 |
| **Total IgE (N=398) Median (95%CI) kU/L** | 468.2 (178.0-828.7) | 330.5 (271.5-410.7) | 1.0001 (0.9998-1.0003) | 0.61 |

N=number; Adj. OR=adjusted odds ratio; CI=confidence interval; SD=standard deviation; m=missing; Columns 2 and 3 represent number (%). \*Adjusted for child's age, sex, area of residence at birth and father's education. Test for trend p-value: <sup>a</sup>=0.08; <sup>b</sup>=0.004; <sup>c</sup>=0.006; <sup>d</sup>=0.02; <sup>e</sup>=0.005. <sup>§</sup>ImmunoCAP® cut-off ≥0.35kU<sub>A</sub>/L.

**Supplementary Table 2:** Risk factors for currently having any of rhinitis, conjunctivitis, eczema and asthma among Ugandan schoolchildren (N=1,693)

| Characteristics | Allergy-related disease in last 12-months |  | Adj. OR<sup>‡</sup> (95% CI) | P-value |
| --- | --- | --- | --- | --- |
|  | Yes (N=734) | No (N=959) |  |  |
| **Age, years** Mean (SD) | 11.34 | 10.91 | 1.05 (1.01-1.09) | 0.01 |
| **Girls** (942) | 388 (52.9) | 554 (57.8) | 0.83 (0.66-1.03) | 0.09 |
| **Father's history of allergic disease** [m=150] |  |  |  |  |
| Yes (363) | 217 (32.2) | 146 (16.8) | 2.06 (1.57-2.70) | <0.0001 |
| **Mother's history of allergic disease** [m=128] |  |  |  |  |
| Yes (592) | 312 (45.3) | 280 (31.9) | 1.66 (1.31-2.11) | <0.0001 |
| **Area of residence at the time of the child's birth** [m=1] |  |  |  |  |
| Rural (355) | 117 (16.0) | 238 (24.8) | 1 |  |
| Town (1,188) | 531 (72.4) | 657 (68.5) | 1.32 (0.99-1.77) |  |
| City (149) | 85 (11.6) | 64 (6.7) | 1.91 (1.22-2.99)<sup>a</sup> | 0.01 |
| **Child's area of residence in first five years of life** [m=1] |  |  |  |  |
| Rural (339) | 110 (15.0) | 229 (23.9) | 1 |  |
| Town (1,267) | 570 (77.8) | 697 (72.7) | 1.41 (1.04-1.89) |  |
| City (86) | 53 (7.2) | 33 (3.4) | 1.78 (1.03-3.06)<sup>b</sup> | 0.04 |
| **Exposure to animals in first five years of life** [m=60] |  |  |  |  |
| Yes (615) | 276 (38.8) | 339 (36.8) | 1.45 (1.14-1.84) | 0.002 |
| **Frequency of 'trucks passing on street near child's home' at enrolment** [m=1] |  |  |  |  |
| Rarely (945) | 370 (50.5) | 575 (60.0) | 1 |  |
| Frequently (624) | 287 (39.1) | 337 (35.1) | 1.18 (0.93-1.49) |  |
| Almost all the time (123) | 76 (10.4) | 47 (4.9) | 2.15 (1.38-3.35)<sup>c</sup> | 0.003 |
| **Use of de-worming medication in last 12 months** [m=3] |  |  |  |  |
| None (578) | 210 (28.7) | 368 (38.4) | 1 |  |
| Once (732) | 304 (41.6) | 428 (44.6) | 1.16 (0.89-1.50) |  |
| ≥Twice (380) | 217 (29.7) | 163 (17.0) | 1.87 (1.38-2.53)<sup>d</sup> | 0.0002 |
| **Any helminth infection at enrolment** [m=130] |  |  |  |  |
| Yes (212) | 78 (11.5) | 134 (15.1) | 0.87 (0.61-1.24) | 0.45 |
| **Skin Prick Test response to any of 7 allergens at enrolment** [m=36] |  |  |  |  |
| Positive (≥3mm) (657) | 364 (50.6) | 293 (31.3) | 1.70 (1.35-2.14) | <0.0001 |
| **Fractional exhaled nitric oxide levels at enrolment** [m=80] |  |  |  |  |
| Elevated (≥35ppb) (381) | 238 (33.2) | 143 (16.0) | 1.99 (1.53-2.59) | <0.0001 |
| **Allergen-specific IgE at enrolment** (N=398)<sup>§</sup> |  |  |  |  |
| Atopic (≥0.35 kU<sub>A</sub>/L) (252) | 159 (71.3) | 93 (53.1) | 1.85 (1.14-3.01) | 0.01 |
| **Total IgE** (N=398) Median (95%CI) kU/L | 432.4 (306.5-595.7) | 274.9 (207.4-339.7) | 1.0001 (0.9999-1.0003) | 0.23 |

N=number; Adj. OR=adjusted odds ratio; CI=confidence interval; SD=standard deviation; m=missing; Columns 2 and 3 represent number (%), unless stated otherwise. <sup>‡</sup>Adjusted for child's age, sex, area of residence at birth and father's education. Test for trend p-value: <sup>a</sup>=0.004; <sup>b</sup>=0.01; <sup>c</sup>=0.002; <sup>d</sup><0.0001. <sup>§</sup>ImmunoCAP<sup>®</sup> cut-off ≥0.35 kU<sub>A</sub>/L.

**Supplementary Table 3:** Risk factors for having ever had any of rhinitis, conjunctivitis and eczema among Ugandan schoolchildren without asthma (N=1,132)

| Characteristics | Allergy-related disease ever |  | Adj. OR<sup>‡</sup> (95% CI) | P-value |
| --- | --- | --- | --- | --- |
|  | Yes (N=634) | No (N=498) |  |  |
| **Age, years** Mean (SD) | 11.19 | 10.64 | 1.05 (1.01-1.09) | 0.02 |
| **Girls** (646) | 384 (60.6) | 262 (52.6) | 1.37 (1.07-1.75) | 0.01 |
| **Father's education** [m=17] |  |  |  |  |
| None/Primary (338) | 183 (29.2) | 155 (31.8) | 1 |  |
| Secondary (401) | 231 (36.8) | 170 (34.8) | 1.13 (0.84-1.51) |  |
| Tertiary (376) | 213 (34.0) | 163 (33.4) | 1.16 (0.32-1.00)<sup>a</sup> | 0.61 |
| **Father's history of allergic disease** [m=113] |  |  |  |  |
| Yes (188) | 138 (24.0) | 50 (11.2) | 2.44 (1.71-3.48) | <0.0001 |
| **Mother's history of allergic disease** [m=95] |  |  |  |  |
| Yes (346) | 237 (40.1) | 109 (24.4) | 2.10 (1.59-2.77) | <0.0001 |
| **Area of residence at the time of the child's birth** |  |  |  |  |
| Rural (280) | 159 (25.1) | 121 (24.3) | 1 |  |
| Town (772) | 419 (66.1) | 353 (70.9) | 0.96 (0.72-1.27) |  |
| City (80) | 56 (8.8) | 24 (4.8) | 1.88 (1.09-3.25)<sup>b</sup> | 0.02 |
| **Exposure to animals in first five years of life** [m=46] |  |  |  |  |
| Yes (426) | 265 (43.4) | 161 (33.8) | 1.49 (1.15-1.93) | 0.002 |
| **Frequency of 'trucks passing on street near child's home' at enrolment** |  |  |  |  |
| Rarely (676) | 357 (56.3) | 319 (64.1) | 1 |  |
| Frequently (392) | 232 (36.6) | 160 (32.1) | 1.30 (1.01-1.69) |  |
| Almost all the time (64) | 45 (7.1) | 19 (3.8) | 1.92 (1.08-3.42)<sup>c</sup> | 0.02 |
| **Use of de-worming medication in last 12 months** [m=1] |  |  |  |  |
| None (427) | 233 (36.8) | 194 (39.0) | 1 |  |
| Once (500) | 259 (40.9) | 241 (48.4) | 0.90 (0.69-1.17) |  |
| ≥Twice (204) | 141 (22.3) | 63 (12.6) | 1.91 (1.33-2.74)<sup>d</sup> | 0.0001 |
| **Any helminth infection at enrolment** [m=87] |  |  |  |  |
| Yes (157) | 78 (13.2) | 79 (17.3) | 0.76 (0.53-1.07) | 0.12 |
| **Skin Prick Test response to any of 7 allergens at enrolment** [m=26] |  |  |  |  |
| Positive (≥3mm) (354) | 226 (36.4) | 128 (26.3) | 1.66 (1.27-2.17) | <0.0001 |
| **Fractional exhaled nitric oxide levels at enrolment** [m=66] |  |  |  |  |
| Elevated (≥35ppb) (181) | 121 (19.8) | 60 (13.2) | 1.61 (1.14-2.27) | 0.007 |
| **Allergen-specific IgE at enrolment** (N=199)<sup>§</sup> |  |  |  |  |
| Atopic (≥0.35 kU<sub>A</sub>/L) (109) | 58 (56.3) | 49 (51.0) | 1.20 (0.68-2.14) | 0.53 |
| **Total IgE** (N=398) Median (95%CI) kU/L | 234.8 (178.3-328.5) | 310.0 (187.6-381.5) | 0.9997 (0.9994-1.0000) | 0.06 |

N=number; Adj. OR=adjusted odds ratio; CI=confidence interval; SD=standard deviation; m=missing; Columns 2 and 3 represent number (%). <sup>‡</sup>Adjusted for child's age, sex, area of residence at birth and father's education. Test for trend p-value: <sup>a</sup>=0.76; <sup>b</sup>=0.17; <sup>c</sup>=0.005; <sup>d</sup>=0.006. <sup>§</sup>ImmunoCAP® cut-off ≥0.35 kU<sub>A</sub>/L.

**Supplementary Table 4:** Risk factors for urticarial rash among schoolchildren in Uganda (N=1,691)

| Characteristics | Urticarial rash ever |  | Adj. OR<sup>‡</sup> (95% CI) | P-value |
| --- | --- | --- | --- | --- |
|  | Yes (N=572) | No (N=1,119) |  |  |
| **Age, years Mean (SD)** | 12.01 (3.04) | 10.64 (3.07) | 1.18 (1.13-1.23) | <0.0001 |
| **Girls (942)** | 339 (59.3) | 603 (53.9) | 1.16 (0.91-1.47) | 0.22 |
| **Fathers' highest education attained [m=20]** |  |  |  |  |
| None/Primary (437) | 168 (29.5) | 269 (24.4) | 1 |  |
| Secondary (595) | 195 (34.3) | 400 (36.3) | 0.71 (0.53-0.94) |  |
| Tertiary (639) | 206 (36.2) | 433 (39.3) | 0.79 (0.59-1.05)<sup>a</sup> | 0.06 |
| **Father's history of allergic disease [m=148]** |  |  |  |  |
| Yes (363) | 147 (27.8) | 216 (21.3) | 1.68 (1.25-2.25) | 0.001 |
| **Mother's history of allergic disease [m=128]** |  |  |  |  |
| Yes (592) | 235 (43.8) | 357 (34.7) | 1.58 (1.23-2.02) | <0.0001 |
| **Area of residence at the time of the child's birth** |  |  |  |  |
| Rural (354) | 126 (22.0) | 228 (20.4) | 1 |  |
| Town (1,188) | 395 (69.1) | 793 (70.9) | 1.15 (0.87-1.53) |  |
| City (149) | 51 (8.9) | 98 (8.7) | 1.08 (0.67-1.74)<sup>b</sup> | 0.60 |
| **Exposure to animals in first five years of life [m=58]** |  |  |  |  |
| Yes (615) | 237 (42.9) | 378 (35.0) | 1.29 (1.01-1.66) | 0.04 |
| **Frequency of 'trucks passing on street near child's home' at enrolment** |  |  |  |  |
| Rarely (944) | 292 (51.1) | 652 (58.3) | 1 |  |
| Frequently (624) | 218 (38.1) | 406 (36.3) | 1.19 (0.93-1.52) |  |
| Almost all the time (123) | 62 (10.8) | 61 (5.4) | 2.19 (1.37-3.48)<sup>c</sup> | 0.003 |
| **Use of de-worming medication in last 12months [m=1]** |  |  |  |  |
| None (578) | 181 (31.7) | 397 (35.5) | 1 |  |
| Once (732) | 237 (41.5) | 495 (44.2) | 0.96 (0.74-1.25) |  |
| ≥Twice (380) | 153 (26.8) | 227 (20.3) | 1.40 (1.02-1.92)<sup>d</sup> | 0.04 |
| **Any helminth infection at enrolment [m=129]** |  |  |  |  |
| Yes (212) | 77 (14.7) | 135 (13.0) | 1.19 (0.84-1.67) | 0.33 |
| **Skin Prick Test response to any of 7 allergens at enrolment [m=35]** |  |  |  |  |
| Positive (≥3mm) (657) | 241 (42.9) | 416 (38.0) | 1.26 (0.99-1.61) | 0.06 |
| **Allergen-specific IgE at enrolment (N=397)<sup>§</sup>** |  |  |  |  |
| Atopic (≥0.35 kU<sub>A</sub>/L) (251) | 94 (67.6) | 157 (60.8) | 1.17 (0.70-1.95) | 0.55 |
| **Total IgE at enrolment (N=398)** |  |  |  |  |
| Median (95%CI) kU/L | 421.3 (234.2-596.0) | 323.3 (271.0-384.8) | 1.00 (0.9998-1.0002) | 0.70 |

N=number; Adj. OR=adjusted odds ratio; CI=confidence interval; SD=standard deviation; m=missing; Columns 2 and 3 represent number (%), unless stated otherwise. <sup>‡</sup>Adjusted for child's age, sex, area of residence at birth and father's education. Test for trend p-value: <sup>a</sup>=0.12; <sup>b</sup>=0.48; <sup>c</sup>=0.002; <sup>d</sup>=0.08. <sup>§</sup> ImmunoCAP®, cut-off ≥0.35 kU<sub>A</sub>/L.

